# Patterns and determinants of risky sexual behaviours among high school students in Lomé, Togo: a cross-sectional study

**DOI:** 10.1101/2025.10.20.25338402

**Authors:** Essi Edjodjinam Kpegba-Fiaboe

## Abstract

**Background:** Adolescent sexual and reproductive health remains a major public health concern in sub-Saharan Africa, where early sexual debut, unsafe practices, and limited agency contribute to adverse outcomes. Despite policy efforts, empirical evidence on risky sexual behaviours (RSBs) among in-school adolescents in Togo remains scarce.

**Methods:** A cross-sectional study was conducted between December 2023 and March 2024 among 461 high school students aged 15–23 years selected through multistage sampling. Data were collected using a structured, self-administered questionnaire and analysed with descriptive statistics, bivariate correlations, and multivariate logistic regression. RSB was constructed as a composite variable comprising inconsistent condom use, concurrent partnerships, substance-influenced sex, and casual sexual encounters.

**Results:** Nearly half (47.3%) of respondents had engaged in sexual intercourse, and among these, 75% reported sexual debut before age 18. Over one-third (39%) had ever been involved in overlapping relationships, while only 56.4% reported inconsistent condom use. More than one-quarter (27.5%) described their first sexual experience as coerced. Logistic regression revealed that students in higher grades, those who initiated sex at or after age 18 (AOR = 5.53, 95% CI 1.54–19.85), and those whose first sexual experience was coerced (AOR = 5.85, 95% CI 1.15– 29.72) were significantly more likely to engage in RSBs. Frequent pornography consumption also increased odds of RSB (AOR = 12.49, 95% CI 4.28–36.46).

**Conclusions:** RSBs among high school students in Lomé reflect complex interactions between developmental, relational, and digital factors. The findings highlight critical gaps in sexual health literacy, consent negotiation, and online safety. Strengthening comprehensive sexuality education, integrating digital literacy into school curricula, and reinforcing gender-transformative interventions are essential to promote informed, safe, and equitable adolescent sexual practices in Togo.

## Introduction

Adolescence represents a pivotal developmental stage characterized by rapid biological maturation, evolving cognitive capacities, and expanding social autonomy. It is during this period that individuals begin to consolidate their identity, assert independence, and navigate increasingly complex interpersonal and sexual relationships [1–3]. As adolescents strive for autonomy, they also experience a profound need for social belonging, rendering them highly responsive to peer norms and social expectations [4]. This dynamic tension between individuation and conformity heightens vulnerability to external pressures, such as misinformation, social coercion, and unequal gender or power relations that can manifest in risky sexual behaviours (RSBs) [3,5].

Globally, adolescent sexual and reproductive health (SRH) remains a major public health concern. Despite substantial progress and policy commitments, the prevalence of negative SRH outcomes (e.g., including early sexual debut, unintended pregnancy, sexually transmitted infections (STIs), and unsafe abortions) remains unacceptably high, particularly in low-and middle-income countries (LMICs) [6,7]. The World Health Organization [8] estimates that approximately 21 million girls aged 15–19 years become pregnant annually, with nearly half of these pregnancies unintended. In sub-Saharan Africa (SSA), one in four adolescent girls gives birth before the age of 18 [9], often resulting in early school dropout, limited socioeconomic mobility, and intergenerational cycles of poverty and gender inequality [10–12].

Across West Africa, structural and sociocultural determinants such as restrictive gender norms, economic precarity, and limited access to youth-friendly SRH services, continue to drive unsafe sexual practices [10,13]. Early sexual initiation, frequently before age 15, coupled with inconsistent contraceptive use, multiple or concurrent partnerships, and transactional relationships remain common among adolescents [7,14–17]. Moreover, silence around sexuality in families and schools restricts open communication, leaving young people to rely on peers or digital media, often sources of misinformation [18–20].

In Togo, national data reflect these regional trends. The Ministry of Health reports that 17% of Togolese girls aged 15–19 have been pregnant or have given birth, with the highest prevalence in the Kara (25.3%) and Maritime (19.0%) regions [21]. According the Togolese Ministry of Education, over 8,600 pregnancies were recorded among pre-university students nationwide (2020–2023), including 2,025 among senior high school girls [22]. Local studies confirm high rates of early sexual initiation and low contraceptive uptake among adolescents [23–25]. Limited parent–child communication around sexuality [26–28] and the stigmatization of contraceptive use further exacerbate these outcomes [23,29,30], leading to unplanned pregnancies, unsafe abortions, and serious reproductive health complications such as haemorrhage, sepsis, and, in severe cases, maternal death [31].

Beyond the risk of pregnancy and its social consequences, RSBs contribute to a significant burden of sexually transmitted infections, including HIV, among adolescents [32,33]. Yet, empirical research in Togo has predominantly focused on university students or out-of-school youth, leaving high school students, a group navigating the intersection of puberty, peer influence, and emerging digital exposure, largely understudied [34,35]. Moreover, existing literature tends to emphasize outcomes rather than determinants, limiting understanding of the behavioural and contextual factors shaping adolescent sexuality in this setting.

The present study addresses this critical gap by examining the patterns and determinants of risky sexual behaviours among high school students in Lomé, Togo. Specifically, it explores early sexual initiation, sexual partnership dynamics, condom use, substance-influenced sex, and transactional encounters, alongside key sociodemographic and contextual correlates. By elucidating these relationships, this study provides empirical evidence to inform youth-centered interventions and comprehensive sexuality education programs aligned with Sustainable Development Goal (SDG) 3.7, which seeks to ensure universal access to sexual and reproductive health information, education, and services by 2030.

## Methods

### Study design and setting

This study employed a cross-sectional quantitative design and was conducted between December 4^th^, 2023 and March 30^th^, 2024 in five high schools in Lomé, Togo, the country’s capital and largest urban center. The selected schools included both public and private institutions, ensuring representation across varying socioeconomic and educational contexts. Lomé was chosen as the study area because of its dense adolescent population, growing digital connectivity, and diversity in school types, which together provide a representative picture of urban adolescent experiences.

### Eligibility criteria

Eligible participants were students aged 15–24 years who were enrolled in one of the selected high schools during the study period. Inclusion criteria required that participants (i) understood French or at least one local language, (ii) were capable of providing informed consent and coherent responses, and (iii) voluntarily agreed to participate in the survey. Students who were outside the target age range, not enrolled in the selected institutions, or unable/unwilling to participate meaningfully were excluded.

### Sample size determination

The minimum required sample size was calculated using Cochran’s formula [36] for cross-sectional studies with an unknown population size:

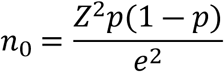

where:

𝑍 = 1.96 corresponds to a 95% confidence level,

𝑝 = 0.5 (assumed proportion of RSB among the study participants), and

𝑒 = 0.05 (margin of error).

The initial sample size obtained was 384. To account for potential non-response or incomplete data (20%), the final sample size was adjusted to 461 participants, which was achieved during data collection.

### Data collection instruments and procedure

A multistage sampling approach was employed for this study. In the first stage, five high schools in Lomé were purposively selected based on size, type (public/private), and accessibility to ensure socioeconomic and institutional diversity. In the second stage, a proportionate stratified sampling method determined the number of participants from each school according to student enrolment. Within each school, simple random sampling was used to select individual respondents from class registers, ensuring balanced representation by sex and grade level (10^th^ –12^th^).

Data were collected using a structured, self-administered questionnaire adapted from validated adolescent SRH surveys [37–39]. The instrument comprised four main sections, each designed to capture different dimensions of SRH behaviours and contexts of young people. The socio-demographic characteristics section collected background information such as age, sex, grade, religion, living arrangement, number of agemates in the household, participation in paid casual work, and mobile device ownership. These variables were used to explore social and structural factors that may shape sexual decision-making of adolescents. In sexual initiation and first sexual experience section, participants were asked whether they had ever engaged in sexual intercourse (“Have you ever had sexual intercourse?” Yes/No) and at what age this first occurred. The nature of the first sexual experience was also assessed using categories such as *willing*, or *coerced (unwilling)*. These questions aimed to identify the timing and consent dynamics surrounding sexual debut.

The third section on sexual partnerships and relationship patterns focused on the number of partners in the past 12 months, and the presence of overlapping (concurrent) sexual relationships. Additional questions examined age disparity with the most recent sexual partner (same age, 3–5 years older, ≥10 years older) and engagement in transactional sex (exchange of money, gifts, or material items for sex). Finally, sexual practices and contexts of sexual activity section captured protective and risk behaviours, including condom use frequency (“always,” “sometimes,” “never”), reasons for inconsistent use, sexual activity under the influence of alcohol or drugs, and experiences of casual or one-time sexual encounters or overlapping relationship. Participants were also asked about exposure to pornography to contextualize broader behavioural pattern associated with sexual risk-taking. The questionnaire was pre-tested among 30 students from a non-participating school in Lomé to evaluate clarity, comprehension, and cultural sensitivity. Feedback from this pilot test informed revisions to wording and structure to improve reliability and response accuracy.

Four trained research assistants fluent in French and local languages (Ewé and Kabyè) administered the questionnaires during scheduled school hours. The survey was conducted in classrooms, with participants completing the anonymous forms individually under supervision to ensure privacy and minimize peer influence. Respondents handed the completed questionnaires in sealed envelopes directly to the researchers to maintain confidentiality. All filled questionnaires were checked daily for completeness and consistency before data entry.

In this study, RSB, the primary outcome variable, was measured using a composite dependent variable comprising four key behavioural indicators: (1) overlapping or concurrent sexual partnerships, (2) inconsistent condom use, (3) sexual activity under the influence of substances, and (4) casual (one-time) sexual encounters. Participants who reported engaging in at least one of these behaviours were categorized as exhibiting “any risky sexual behaviour”, resulting in a binary outcome variable coded as *0 = No* and *1 = Yes*. This dependent variable was tested for association with socio-demographic factors, including age group, sex, grade level, religion, living arrangement, number of agemates in the household, engagement in paid work, and mobile device ownership. Additionally, behavioural and contextual factors, such as age at sexual debut, nature of first sexual experience, number of sexual partners in the past 12 months, partner age difference, engagement in transactional sex, and exposure to pornography, were also analyzed to account for variations in sexual behaviour patterns and relational context.

### Data analysis

Data was entered, cleaned and coded in Excel spreadsheet and then analyzed using Stata (version 17). Descriptive statistics (frequencies, percentages, means, and standard deviations) summarized participants’ characteristics and sexual behaviour patterns. Binary logistic regression was used to examine the association between RSB and the independent variables. Bivariate logistic regression analyses were first conducted to explore the relationship between each independent variable and RSB, followed by multivariable logistic regression to identify independent predictors of risky sexual behavior. Both Crude Odds Ratios (COR) and Adjusted Odds Ratios (AOR) with 95% confidence intervals were calculated to determine the strength and direction of associations. Statistical significance was set at *p* < 0.05.

### Ethical considerations

The study was conducted in full adherence to national and international ethical standards for research involving human participants. Ethical approval was obtained from the National Ethics Committee for Health Research in Togo (Protocol No. 047/2023/CBRS) and the Ethics Committee for the Humanities, University of Ghana (Approval No. ECH 285/22-23). Additional authorization was granted by the Ministry of Primary, Secondary, and Technical Education in Togo (Ref. 2454-23/DRE-GL), the Regional Directorate of Education, and the respective school administrations. Written informed consent was obtained from all participants aged 18 years and above, while assent and parental consent were obtained for those under 18. To protect participants’ privacy and confidentiality, each respondent was assigned a unique identification code; no names or identifiable details were recorded. Data were securely stored in password-protected files accessible only to the research team. Although the study posed no foreseeable risks, a trained mental health professional was available during data collection to provide immediate support to participants who might have experienced distress while responding to questions on sensitive sexual and reproductive health topics.

## Results

### Socio-demographic characteristics of the study’s participants

A total of 461 high school students participated in the study (Table 1). Slightly more than half of the respondents were male (53.8%), while females accounted for 46.2%. The majority (89.1%) were aged between 15 and 18 years, with a mean age of 16.9 years (SD = 2.0). In terms of grade level, 37.5% were in 10^th^ grade, 33.6% in 11^th^ grade, and 28.9% in 12^th^ grade. Regarding household composition, about 62% reported living with more than three agemates, while 22% lived with one or two, and 16% had no agemates at home. Christianity was the predominant religion among respondents (47.7%), followed by Islam (41.2%), while 11.1% reported as animists or having no religious affiliation. Nearly half (47.5%) of participants lived with both parents, while 37.7% lived with a single parent, 11.7% with other relatives or non-relatives, and 3.1% reported living alone. In terms of economic engagement, the majority (69.4%) had never engaged in paid casual work, 19.1% did so occasionally, and 11.5% reported working often. Concerning mobile device ownership, 41.4% of respondents primarily accessed a peer’s device (shared phone use), while 58.6% owned a personal mobile device.

**Table 1.** Socio-demographic characteristics of the participants.

| Variable | Category | Frequency<br>(N = 461) | Percentage |
| --- | --- | --- | --- |
| Sex | Male | 248 | 53.8 |
|  | Female | 213 | 46.2 |
| Age group | 15–18 | 411 | 89.1 |
|  | 19–23 | 50 | 10.9 |
| Mean age (±SD) | 16.9±2.0 | – | – |

| Variable | Category | Frequency<br>(*N* = 461) | Percentage |
| --- | --- | --- | --- |
| **Grade** | 10<sup>th</sup> Grade | 173 | 37.5 |
|  | 11<sup>th</sup> Grade | 155 | 33.6 |
|  | 12<sup>th</sup> Grade | 133 | 28.9 |
| **Agemates in household <sup>a</sup>** | None | 73 | 15.8 |
|  | 1–2 | 103 | 22.4 |
|  | 3–4 | 184 | 39.9 |
|  | 5+ | 101 | 21.9 |
| **Religion** | Christianity | 220 | 47.7 |
|  | Islam | 190 | 41.2 |
|  | Other religions <sup>b</sup> | 41 | 11.1 |
| **Living arrangement** | Both parents | 219 | 47.5 |
|  | Single parent | 174 | 37.7 |
|  | Other relative/non-relative | 54 | 11.7 |
|  | Living alone | 14 | 3.1 |
| **Paid casual work** | Never | 320 | 69.4 |
|  | Occasionally | 88 | 19.1 |
|  | Often | 53 | 11.5 |
| **Mobile device ownership** | Peer's device <sup>c</sup> | 191 | 41.4 |
|  | Personal device | 270 | 58.6 |

<sup>a</sup> Agemates in household refer to other individuals of similar age living in the same household.
<sup>b</sup> “Other religion” includes respondents who identified with traditional beliefs (e.g., Animism) or reported having no religious affiliation.
<sup>c</sup> Peer's device refers to a mobile phone belonging to a friend or classmate that the respondent uses.

### Patterns of sexual behaviour among high school students

Out of the total sample of 461 respondents, nearly half (47.3%) reported having ever engaged in sexual intercourse, while 52.7% indicated that they had never done so (Table 2). Among those who were sexually active (n = 218), more than half (58.7%) reported initiating sexual activity before the age of 18, whereas 41.3% experienced sexual debut at 18 years or older.

**Table 2.**
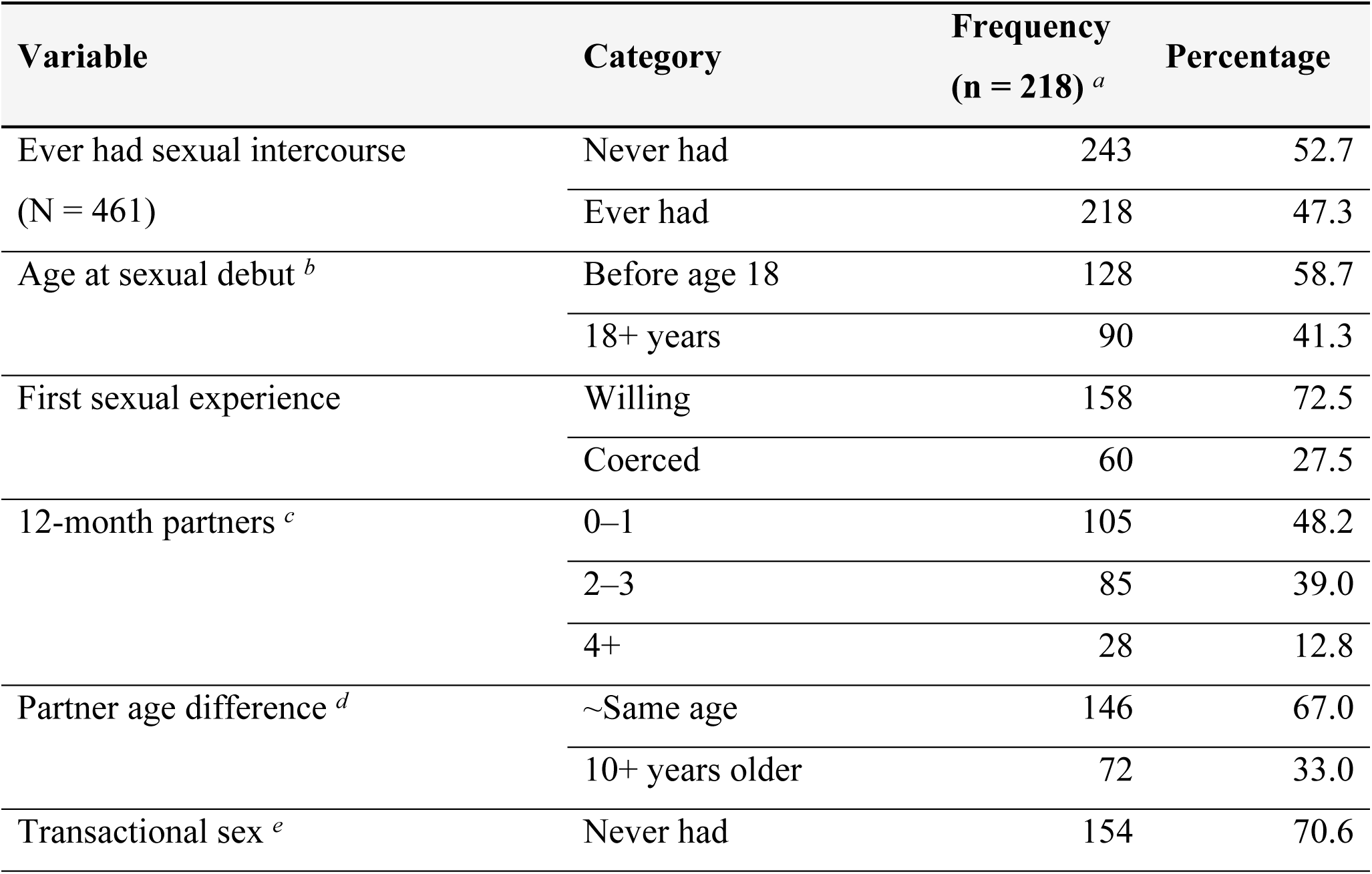
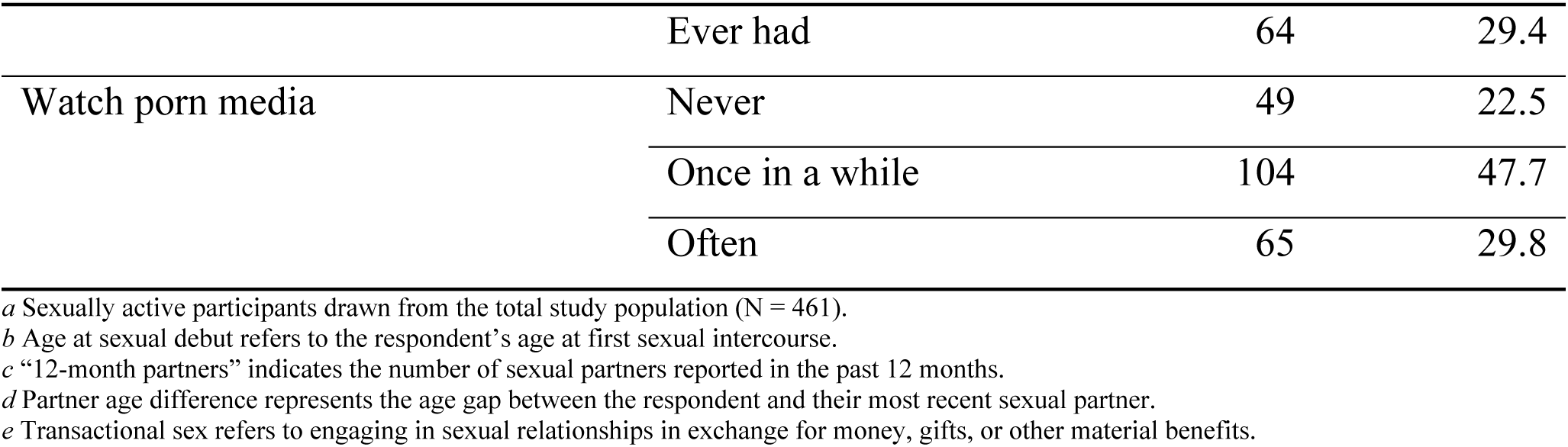
Partner characteristics and sexual relationships.

Regarding the nature of first sexual experience, the majority (72.5%) described it as willing, while 27.5% reported being coerced. In terms of recent sexual activity, almost half of respondents (48.2%) reported having at most one sexual partner in the past 12 months, 39% had two to three partners, and 12.8% had four or more partners during the same period. With respect to partner age difference, two-thirds (67%) reported that their most recent partner was of roughly the same age, while one-third (33%) engaged with partners who were ten or more years older. Nearly three in ten respondents (29.4%) reported having engaged in transactional sex, involving the exchange of money, gifts, or other material benefits. Regarding exposure to sexually explicit content, almost half of the respondents (47.7%) indicated watching pornographic media once in a while, 29.8% reported frequent viewing, and 22.5% stated they had never watched such content.

### Risky sexual behavior (RSB) among sexually-active high school students

Among the sexually active participants (n = 218), 169 students (77.5%) reported engagement in at least one form of RSB (Table 3). Over one-third (39%) indicated having ever been involved in overlapping or concurrent sexual relationships, while 59% reported no such experience (Figure 1A). In relation to condom use, only 43.6% of respondents reported consistent use during sexual intercourse, whereas nearly half (46.3%) used condoms only sometimes, and 10.1% stated that they never used them (Figure 1B). Regarding substance-influenced sexual activity, the majority (89%) reported never having engaged in sex under the influence of alcohol or drugs, while 11% indicated that they had done so (Figure 1C). Furthermore, approximately one in five respondents (20.6%) reported having engaged in occasional or one-time sexual intercourse, with 15.1% having done so once and 5.5% having engaged in such encounters on two or more occasions (Figure 1D).

**Figure 1.**
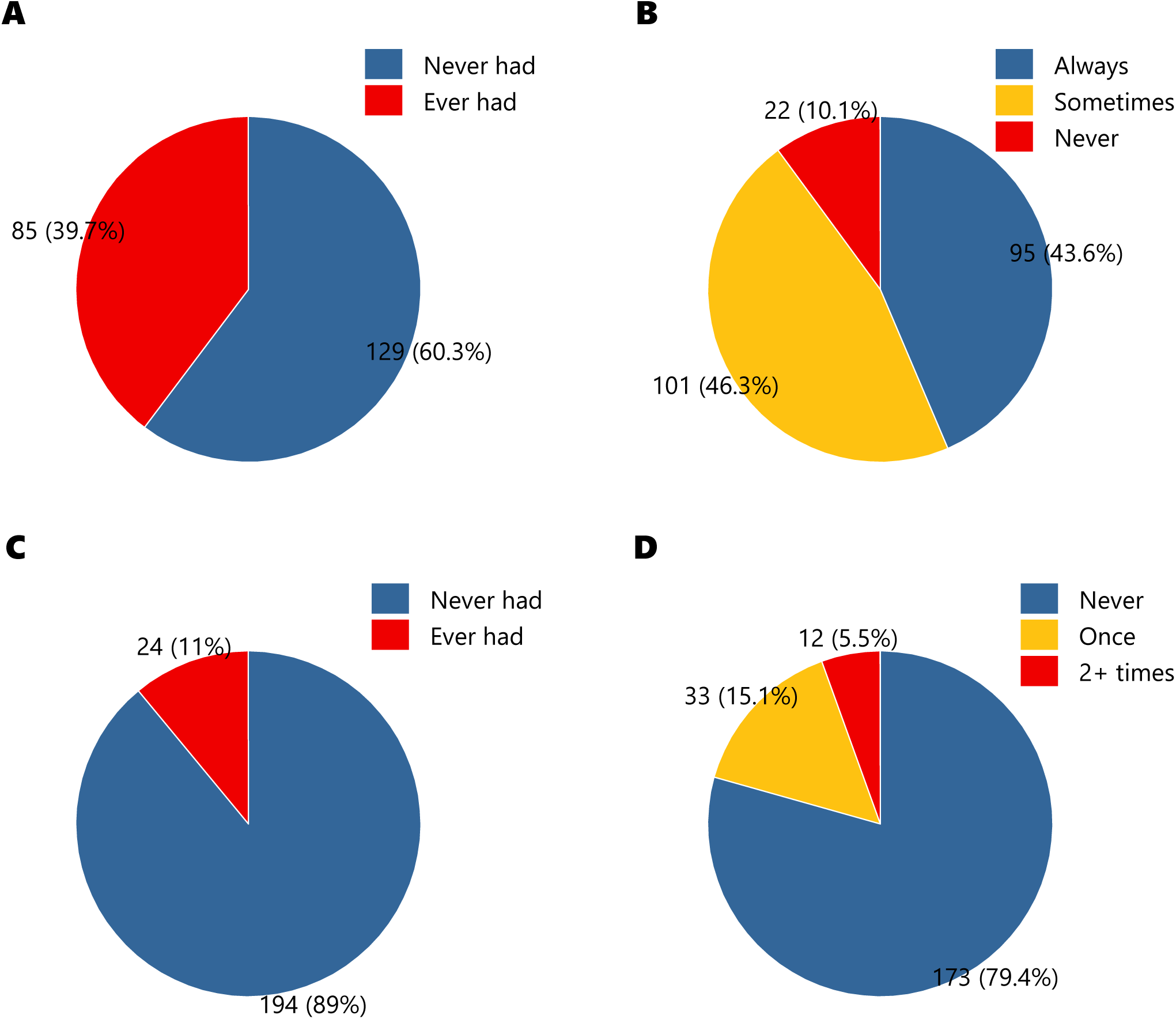
Patterns of risky sexual behaviours among high school students in Lomé, Togo (2024). Distribution of overlapping sexual relationships (**A**), condom use consistency (**B**), substance-influenced sexual activity (**C**), and casual (one-time) sexual encounters (**D**) among sexually active respondents (n = 218).

**Table 3.** Determinants of risky sexual behavior (RSB) among sexually-active high school students in Lomé, Togo, 2024 (n = 218).

| Variable | RSB n (%) |  | COR<br>(95% CI) | AOR<br>(95% CI) |
| --- | --- | --- | --- | --- |
|  | No<br>(n = 49) | Yes<br>(n = 169) |  |  |
| Sex <sup>[Male]</sup> | 22 (21.6) | 80 (78.4) | 1 |  |
|  | No<br>(n = 49) | Yes<br>(n = 169) |  |  |
| Female | 27 (23.3) | 89 (76.7) | 0.90<br>(0.47–1.71) | 0.42<br>(0.11–1.55) |
| **Age** <sup>[15–18]</sup> | 46 (24.7) | 140 (75.3) | 1 |  |
| 19 – 23 | 3 (9.4) | 29 (90.6) | 3.17<br>(0.92–10.91) | 2.16<br>(0.29–16.12) |
| **Grade** <sup>[10th]</sup> | 23 (34.8) | 43 (65.2) | 1 |  |
| 11<sup>th</sup> | 15 (20.0) | 60 (80.0) | 2.13<br>(1.001–4.57) | 1.88<br>(0.55–6.37) |
| 12<sup>th</sup> | 11 (14.3) | 66 (85.7) | 3.20**<br>(1.42–7.24) | 1.32<br>(0.36–4.85) |
| **Agemates in household** <sup>[None]</sup> | 5 (9.8) | 46 (90.2) | 1 |  |
| 1–2 | 24 (27.3) | 64 (72.7) | 0.28*<br>(0.10–0.81) | 0.23<br>(0.55–1.00) |
| 3–4 | 14 (24.6) | 43 (75.4) | 0.33<br>(0.11–1.01) | 0.41<br>(0.09–1.86) |
| 5+ | 6 (27.3) | 16 (72.7) | 0.28<br>(0.77–1.08) | 0.57<br>(0.09–3.63) |
| **Religion** <sup>[Christianism]</sup> | 38 (20.0) | 152 (80.0) | 1 |  |
| Islam | 5 (33.3) | 10 (66.7) | 0.49<br>(0.16–1.54) | 0.44<br>(0.88–2.26) |
| Other religions | 6 (46.2) | 7 (53.8) | 0.29*<br>(0.09–0.91) | 0.07**<br>(0.01–0.37) |
| **Living arrangement** <sup>[Both parent]</sup> | 25 (25.5) | 73 (74.5) | 1 |  |
| Single parent (Mother/Father) | 11 (13.1) | 73 (86.9) | 2.27*<br>(1.04–4.95) | 2.11<br>(0.65–6.85) |
| Other relative/non relative | 8 (34.8) | 15 (65.2) | 0.64<br>(0.24–1.69) | 0.31<br>(0.07–1.37) |
| Alone | 5 (38.5) | 8 (61.5) | 0.54<br>(0.16–1.83) | 0.04**<br>(0.00–0.40) |
| **Paid casual work** <sup>[Never]</sup> | 35 (25.0) | 105 (75.0) | 1 |  |
| Occasionally | 9 (18.0) | 41 (82.0) | 1.51<br>(0.67–3.43) | 1.16<br>(0.33–4.06) |
| Often (at least once a month) | 5 (17.9) | 23 (82.1) | 1.53<br>(0.54–4.33) | 0.86<br>(0.21–3.46) |
| **Mobile device ownership** <sup>[Do not own]</sup> | 39 (44.8) | 48 (55.2) | 1 |  |
| Personal phone | 10 (7.6) | 121 (92.4) | 9.83***<br>(4.54–21.25) | 11.35***<br>(3.11–41.40) |
| **Age at sexual debut** <sup>[Before age 18]</sup> | 40 (31.3) | 88 (68.7) | 1 |  |
| 18+ years | 9 (10.0) | 81 (90.0) | 4.09***<br>(1.86–8.95) | 5.53**<br>(1.54–19.85) |
|  | No<br>(n = 49) | Yes<br>(n = 169) |  |  |
| **First sexual experience** <sup>[Willing]</sup> | 45 (28.5) | 113 (71.5) | 1 |  |
| Coerced | 4 (6.7) | 56 (93.3) | 5.57**<br>(1.90–16.27) | 5.85*<br>(1.15–29.72) |
| **12-month partners** <sup>[0–1]</sup> | 33 (31.4) | 72 (68.6) | 1 |  |
| 2–3 | 10 (11.8) | 75 (88.2) | 3.43**<br>(1.57–7.48) | 2.10<br>(0.66–6.65) |
| 4+ | 6 (21.4) | 22 (78.6) | 1.68<br>(0.62–4.53) | 2.15<br>(0.41–11.15) |
| **Partner age difference** <sup>[~Same age]</sup> | 37 (25.3) | 109 (74.7) | 1 |  |
| 10+ years older | 12 (16.7) | 60 (83.3) | 1.69<br>(0.82–3.49) | 0.99<br>(0.29–3.37) |
| **Transactional sex** <sup>[No]</sup> | 36 (23.4) | 118 (76.6) | 1 |  |
| Yes | 13 (20.3) | 51 (79.7) | 1.19<br>(0.58–2.44) | 0.82<br>(0.24–2.74) |
| **Pornography** <sup>[Never]</sup> | 25 (51.0) | 24 (49.0) | 1 |  |
| Once in a while | 19 (18.3) | 85 (81.7) | 4.66***<br>(2.20–9.85) | 2.50<br>(0.78–7.94) |
| Often | 5 (7.7) | 60 (92.3) | 12.49***<br>(4.28–36.46) | 4.79<br>(0.82–27.88) |

<sup>a</sup> [Superscript] indicates reference categories for comparison.
\* $p < 0.05$ ; \*\* $p < 0.01$ ; \*\*\* $p < 0.001$ .
COR = Crude Odds Ratio; AOR = Adjusted Odds Ratio; CI = Confidence Interval.

### Determinants of risky sexual behaviour among high school students

In the bivariate model, students in higher grades were more likely to report RSB compared to those in 10^th^ grade (COR = 3.20; 95% CI 1.42–7.24; *p* = 0.005; Table 3). Having at least one or two agemates in the household was significantly associated with lower odds of RSB compared to those without agemates (COR = 0.28; 95% CI 0.10–0.81; *p* = 0.019).

Multivariate logistic regression analyses showed that religious affiliation, living arrangement, sexual history, ownership of digital device, and pornography showed a relationship with RSB. Respondents identifying with other religions had significantly lower odds of engaging in RSB than Christians (AOR = 0.07; 95% CI 0.01–0.37; *p* = 0.002). Similarly, adolescents living alone had markedly lower odds of RSB compared to those living with both parents (AOR = 0.04; 95% CI 0.00–0.40; *p* = 0.006). Access to digital technology emerged as a strong predictor. Participants who owned a personal mobile phone had over eleven times higher odds of engaging in RSBs than those without one (AOR = 11.35; 95% CI 3.11–41.40; *p* < 0.0001).

Regarding sexual history, those who initiated sex at or after age 18 were significantly more likely to engage in RSBs (AOR = 5.53; 95% CI 1.54–19.85; *p* = 0.009), as were individuals whose first sexual experience was coerced (AOR = 5.85; 95% CI 1.15–29.72; *p* = 0.033). Similarly, participants who had two to three sexual partners in the 12 months preceding data collection were significantly more likely to engage in RSB compared to those who had only one sexual partner (COR = 3.43; 95% CI 1.57–7.48; *p* = 0.002). Pornography consumption also demonstrated a strong association. Adolescents who frequently watched pornographic material were substantially more likely to engage in RSB than those who never did (COR = 12.49; 95% CI 4.28–36.46; *p* < 0.0001). Other variables, including age, transactional sex, and partner age difference, were not significantly associated with RSB in the adjusted model (*p* > 0.05; Table 3).

## Discussion

This study examined the patterns and determinants of RSBs among high school students in Lomé, Togo, providing important insights into the behavioural and contextual dynamics that shape adolescent sexuality in an urban West African context. Nearly half of the respondents reported having initiated sexual activity, a prevalence consistent with findings from other urban populations across SSA, where sexual debut before the age of 18 remains widespread [7,40,41].

Early sexual initiation, reported by nearly three-quarters of sexually active participants, is particularly worrisome given its well-established association with increased vulnerability to unprotected intercourse and STIs [14,42,43]. Intriguingly, however, adolescents who initiated sex at or after age 18 were significantly more likely to engage in RSBs compared to those who debuted earlier. This seemingly paradoxical finding may overlap with schooling progression: older adolescents, typically in higher grades, experience greater social freedom, exposure to peer and romantic networks, and reduced parental supervision [44]. Consistent with this interpretation, grade level was also significantly associated with RSB, with students in higher grades more likely to report risky behaviours compared to those in 10^th^ grade. This pattern aligns with evidence from longitudinal school-based studies, which consistently demonstrate that as students advance through grades, they are exposed to greater opportunities for experimentation and risk-taking behaviours, including substance use and sexual activity, often accompanied by a reduced perception of potential harm [45–47]. Together, these findings suggest that academic advancement often coincides with expanded social opportunities and experimentation, yet without a corresponding improvement in sexual health literacy. Formal education alone may not translate into informed sexual decision-making, leaving many young people “educationally advanced but sexually uninformed.” Consequently, delayed sexual debut among older students may not signify abstinence based on informed choice but rather postponed exposure that, once initiated, occurs with inadequate preparation or limited risk awareness. This dynamic may manifest in impulsive or unprotected encounters, overlapping sexual relationships, or contextually pressured sexual activity [47–49].

Notably, more than half of the sexually active participants in this study reported having had multiple sexual partners in the 12 months preceding data collection, and two in five acknowledged overlapping relationships. These patterns are concerning, as multiple and concurrent partnerships substantially increase both the frequency and diversity of sexual exposures, thereby elevating the risk of unintended pregnancies, as well as STIs, including HIV and trichomoniasis [45,50–52]. Such relationships often occur within contexts marked by inconsistent condom use, emotional instability, and unequal power dynamics, further exacerbating adolescents’ vulnerability to infection and reproductive health complications [10,53,54]. These findings reinforce the urgent need for targeted interventions that address not only behavioural risk but also the underlying relational and psychosocial determinants that sustain high-risk sexual networks among young people.

In addition to behavioural experimentation, relational power dynamics emerged as a critical determinant of adolescent sexual risk. More than one-quarter of participants described their first sexual experience as coerced, and these individuals were nearly six times more likely to engage in subsequent RSBs. This association reinforces the long-recognized link between early exposure to sexual coercion and diminished sexual agency, with enduring consequences for consent negotiation and protective behaviour [55,56]. The lingering psychosocial impacts of coercion (such as internalized disempowerment, lowered self-worth, and normalization of non-consensual experiences) can entrench cycles of vulnerability, as supported by prior studies [57,58]. These findings highlight the urgent need for comprehensive, gender-transformative sexuality education that extends beyond biological instruction to include critical discussions on consent, power dynamics, and emotional wellbeing [59]. Embedding these themes in both school curricula and community-based programs can strengthen adolescents’ capacity to recognize, resist, and report coercive encounters while promoting respectful, equitable relationships.

Despite Togo’s legal frameworks, such as the *Code de l’Enfant* (Law No. 2007–017) and commitments under the *African Charter on the Rights and Welfare of the Child* [60], the persistence of coercive first sexual experiences underscores gaps in implementation and entrenched sociocultural resistance to gender equity [61,62]. Strengthening the enforcement of these protections through confidential school-based reporting systems, youth advocacy networks, and community sensitization initiatives remains essential for safeguarding adolescents’ sexual and reproductive rights.

Beyond interpersonal and contextual determinants, this study also highlights the emerging influence of digital exposure in shaping adolescent sexual behaviours. In an era where online connectivity increasingly mediates social interactions, the digital ecosystem functions as both an enabler of information access and a potential catalyst for sexual experimentation and risk. Findings from this study revealed that adolescents who owned personal mobile devices were over eleven times more likely to engage in RSBs compared to those who do not or with inconsistent access. This striking association suggests that private, unregulated access to smartphones may expand adolescents’ opportunities for sexual networking, exposure to explicit content, and unsupervised digital interactions, often without corresponding guidance or digital literacy [19,63].

While digital platforms can facilitate access to credible SRH information, they simultaneously expose users to sexually explicit material, peer pressure, and online solicitation [64,65]. In the present study, nearly one in three respondents reported frequent consumption of pornographic content, an activity that has been consistently linked to distorted sexual norms, objectification, and increased likelihood of multiple partnerships or unprotected sex [41,66,67]. Such findings reflect the “digital double bind” of adolescence: while smartphones can empower young people with knowledge, they also serve as unfiltered conduits for sexualized media that normalize RSBs [19,68]. These dynamics highlight the need to reconceptualize SRH promotion for the digital generation.

This “digital double bind” underlines the urgent need for policies and programs that balance access with protection in the rapidly evolving online landscape. In Togo, where digital connectivity among adolescents is expanding but regulatory and educational systems lag behind, interventions must move beyond content restriction to foster digital resilience, the ability to critically navigate, assess, and respond to online sexual content [69,70]. This requires integrating *digital literacy and online safety education* into secondary school curricula, empowering students to recognize harmful content, resist peer pressure, and make informed decisions about their digital interactions [64].

At the institutional level, partnerships between the Ministry of Education, the Ministry of Digital Economy and Transformation, and youth-led organizations could facilitate the co-development of safe digital learning ecosystems, including moderated online SRH discussion forums, verified educational social media pages, and integration of parental digital guidance modules. For example, expanding the *InfoAdoJeunes* mobile platform (an mHealth platform providing youth-friendly SRH information and counselling), to include interactive digital literacy modules, peer mentorship spaces, and anonymous reporting tools for sexual violence, online harassment could significantly strengthen its preventive and educational reach.

Moreover, media regulation bodies such as the *Haute Autorité de l’Audiovisuel et de la Communication (HAAC)* could collaborate with mobile network operators and youth advocates to promote ethical digital citizenship campaigns that emphasize respectful online behaviour, counter sexual exploitation, and challenge the normalization of coercive sexual narratives. Community-level interventions led by organizations such as the *Association Togolaise pour le Bien-Être Familial (ATBEF)* can further complement these efforts by combining offline dialogues through youth clubs, school debates, and peer-education programs, with online campaigns designed to engage adolescents in reflection and responsible sexual decision-making.

Taken together, these approaches position digital technology not merely as a source of risk, but as a potential ally in strengthening adolescents’ sexual health literacy, agency, and resilience. Building safe, empowering digital environments will require multisectoral collaboration, linking education, technology, health, and youth engagement sectors to ensure that Togo’s digital transition supports, rather than undermines, adolescent wellbeing and reproductive rights.

## Conclusion

This study provides critical insights into the multifaceted determinants of RSBs among high school students in Lomé, Togo. The findings reveal that sexual risk-taking among young people is not a product of isolated decisions but a reflection of broader developmental, relational, and digital influences. Early sexual debut, multiple and concurrent partnerships, and coercive first sexual experiences collectively underscore the persistent gaps in sexual agency, consent negotiation, and health literacy. Meanwhile, the strong association between mobile phone ownership, pornography exposure, and RSBs highlights the emerging challenges of adolescent sexuality in the digital era, where access to information coexists with exposure to harmful and unregulated content. These findings highlight the urgent need for multisectoral, gender-transformative, and digitally responsive interventions that extend beyond traditional classroom-based education. Strengthening comprehensive sexuality education across the secondary school cycle, embedding digital literacy and online safety into SRH curricula, and enforcing national child protection frameworks can together foster safer, more informed, and empowered adolescent sexual behaviours. Collaborative efforts involving schools, parents, health providers, technology regulators, and community organizations are essential to transform both physical and digital environments into supportive spaces for young people’s sexual and reproductive wellbeing. Ultimately, advancing young people’s SRH in Togo requires shifting from reactive prevention to proactive empowerment, cultivating a generation that is not only knowledgeable but also equipped with the confidence, agency, and critical thinking needed to navigate the complex realities of modern sexuality.

## Data Availability

All relevant data are within the manuscript.

## Acknowledgements

This paper forms part of the author’s doctoral research in Population Studies. The author gratefully acknowledges the German Academic Exchange Service (DAAD) for supporting her PhD training. Deep appreciation is also extended to her supervisors—Prof. Adriana A. Biney, Prof. Stephen O. Kwankye, and Dr. Yaw Atiglo—for their valuable guidance and review of earlier drafts. The author thanks the participating schools and students for their cooperation and contribution to this study.

## Data availability statement

All relevant data are within the manuscript.

